# Classification of Alzheimer’s disease in a mixed clinical cohort using biofluid Raman spectroscopy

**DOI:** 10.1101/2025.02.23.25322672

**Authors:** George Devitt, Sofia K Michopoulou, Latha Kadalayil, Niall Hanrahan, Angus Prosser, Boyd Ghosh, Amrit Mudher, Christopher M Kipps, Sumeet Mahajan

## Abstract

There is a critical unmet need for scalable, accessible and objective diagnostic tests for stratification in dementia. Biofluid Raman spectroscopy (RS) due to its simplicity, holistic and label-free nature, is a powerful approach that has the potential to offer differential diagnosis across dementia types including Alzheimer’s disease (AD). RS is a laser-based optical method that can rapidly provide chemically rich information (‘spectral biomarkers’) from biofluids but its utility for AD diagnosis has not been established in a ‘real-world’ context, specifically from a clinically heterogenous cohort of patients. We carried out RS measurements on cerebrospinal fluid (CSF) samples of patients from a mixed clinical cohort (N=143). All patients reported cognitive complaints and were clinically diagnosed over 2 years with conditions including AD and other neurodegenerative diseases, as well as developmental and long-term chronic conditions. Machine-learning algorithms were trained, optimized and evaluated on Raman spectra to classify AD from non-AD. AD was classified with 93% accuracy for patients in the testing set. Time from sample to classification was < 1 hour. Spectral biomarkers explaining AD classification were identified and primarily assigned to protein-derived aromatic amino acids, representing a difference in proteome signature between AD and non-AD groups. Signals from a subset of spectral biomarkers directly correlated with pathological CSF biomarker concentrations including amyloid-β 42, phosphorylated-tau 181, and total tau. This pre-clinical study is a first step towards realizing the real-world application of RS for dementia diagnosis. Compared to current and emerging methods, RS does not require sophisticated instrumentation or specialized labs. It is reagentless and simple, offering unprecedented rapidity, scalability, accessibility for dementia diagnosis.

## INTRODUCTION

Clinical diagnosis of Alzheimer’s disease (AD) is often inconsistent with post-mortem neuropathology at autopsy, which is necessary for a definitive AD diagnosis ^1–3^. Reported accuracies of clinical AD diagnosis vary widely ^4^, in part due to diagnostic inaccuracies and patient heterogeneity. To address this, there has been a move from a symptomatic definition of AD towards a biological definition based on pathological biomarkers, including amyloid-β (Aβ) and tau protein biofluid levels ^3^. However, there is still a need to develop more accessible and objective biomarkers that are highly sensitive and specific to AD, whilst retaining stratification capability over other clinically overlapping neurodegenerative diseases (NDDs). Such biomarkers could improve AD diagnosis rate and accuracy, as well as patient cohort selection and stratification in drug trials. Furthermore, patient selection for AD disease-modifying therapies like Lecanemab ^5^ underscores the importance of identifying those most likely to benefit and tailoring interventions accordingly.

Current biomarkers for AD include structural and functional imaging such as magnetic resonance imaging (MRI) and single-photon emission computed tomography (SPECT)/positron emission tomography (PET). Biofluid markers are routinely utilized, particularly cerebrospinal fluid (CSF) markers for amyloid, tau and neurodegeneration (ATN), specifically amyloid-β-42 (Aβ42, A) phosphorylated tau at site 181 (p-tau, T), and total tau (t-tau, N). Several novel biomarkers are emerging as candidates to further support AD diagnosis, including plasma p-tau 217 ^3,6^, as well as omics-based (meta-genomics, proteomics, metabolomics) biomarkers ^7^, and tau-specific PET ligands ^8^. A simpler yet powerful solution may be provided by optical spectroscopy. Raman spectroscopy (RS) is an optical technique that can provide highly selective label-free readouts within seconds and has the potential for portability at scale ^9^. RS is label-free and reagentless and can be carried out with a small footprint using portable devices in a range of healthcare settings, potentially without the need of skilled operators. Biofluids can be analysed directly to provide an unbiased and holistic chemical fingerprint based on the vibrational modes of the molecules within the sample. The RS-based chemical fingerprinting approach represents the collective characteristics of the biofluid and contrasts with the detection of a specific molecule or analyte ^10^. Therefore, Raman spectroscopy may have utility in the detection and stratification of multiple, if not all, neurodegenerative diseases, opening the door to rapid differential diagnosis and therefore timely personalized intervention.

Proof-of-concept studies have demonstrated that RS may have clinical utility for the identification of AD (the most common cause of dementia) in CSF, but small patient sample sizes of 19 ^11^ and 37 ^12^ together with the lack of comparative clinical gold standards currently limit any confidence in clinical validity. Additionally, these studies used healthy controls as comparators to AD patients, which does not reflect real-world diagnostic scenarios as it is unlikely that ‘healthy’ people will seek an AD diagnosis. A more appropriate control group will be patients who sought dementia diagnoses due to cognitive complaints but were deemed to have a condition other than AD, such as another neurodegenerative disease. It is therefore still not established whether RS can detect AD in a clinical cohort, representative of the clinical setting or population, with appropriate statistical power.

To address this critical gap, we utilized a mixed clinical cohort of patients (N=143) who reported cognitive complaints and for whom diagnosis was not initially clear. CSF samples, taken by lumbar puncture, were used for ATN biomarker analysis. We hypothesized that RS analysis of CSF can provide unique spectral fingerprints of AD-specific pathology/neurodegeneration enabling differentiation between AD and non-AD patients even in a heterogenous cohort. We tested AD classification of Raman spectra using machine-learning (ML) algorithms. From the Raman spectra we extracted spectral biomarkers specific to AD and assessed their association with CSF ATN biomarkers by correlation. Our results demonstrate an important step towards the clinical translation of Raman spectroscopy for supporting AD diagnosis, and potentially differential dementia diagnosis, in the future.

## MATERIALS AND METHODS

### Study design

The current study utilized 143 CSF samples collected from patients with cognitive complaints referred to Wessex Cognitive Disorders Clinic at University Hospital Southampton NHS Foundation Trust between 2014 and 2021 for diagnostic lumbar puncture to assess neurodegenerative pathology. All samples that had been annotated with patient data including age, sex, clinical diagnosis and CSF biomarker concentrations were collected and used for this study. Inclusion criteria for the current study were referral for cognitive complaints with query of dementia, availability of AD CSF biomarker results, and availability of CSF sample for Raman analysis. Exclusion criteria were not applied. The cohort was divided into two groups including AD and non-AD (Table 1) based on clinical diagnosis that used a combination of clinical test scores, brain imaging, and CSF biomarker results. The research objectives were to classify CSF samples into AD and non-AD groups based on cross-sectional analysis using Raman spectroscopy measurements in combination with machine-learning. The investigator was blinded to patient diagnosis during spectroscopy measurements and data preprocessing and unblinded at the machine-learning stage as all models used as supervised.

**Table 1.** CSF sample cohort.

| Clinical diagnosis | Age (mean ± SD) | Sex (%M / %F) | Number of patients |
| --- | --- | --- | --- |
| Non-AD | 68 ± 11 | 71 / 29 | 76 |
| AD | 64 ± 9 | 43 / 57 | 67 |
| Total | 66 ± 10 | 65 / 35 | 143 |
| Footnotes: CSF (cerebrospinal fluid), SD (standard deviation) AD (Alzheimer's disease) |  |  |  |

### CSF samples

CSF biomarker concentrations for Aβ42, total tau, and p-tau 181 were measured by the UKAS accredited Neuroimmunology and CSF laboratory at the National Hospital for Neurology and Neurosurgery at Queen Square London using the INNOTEST (Fujirebio) enzyme linked immunosorbent assays (ELISA) system until 2020, and the Lumipulse G (Fujirebio) chemiluminescent immunoassay since 2020 (each of which has consistent results ^13^). The AD group included different AD variants (amnestic, speech, visual), as well as AD with other chronic comorbidities. Diagnoses in the non-AD group spanned a range of clinical conditions including dementia caused by other NDDs, as well as developmental and chronic conditions not related to neurodegeneration. A full list of clinical diagnoses can be found in Supplementary Table 1. Samples were collected as part of two studies that were approved by the Research Ethics Committee (REC:20/NW/0222 and REC:15/SC/0231). Written informed consent was provided at the time of the lumbar puncture by participants or their next of kin, including permission for the storage of excess CSF and use in research studies.

### Sample handling and storage

A minimum of 2 ml of CSF was collected from each participant by lumbar puncture and centrifuged within 30 min in Starstedt polypropylene tubes. 500μL were aliquoted in an Elkay polypropylene tube, frozen at –70°C and sent for clinical diagnostic analysis to the Neuroimmunology and CSF laboratory at University College London Hospitals NHS Foundation Trust (UCLH). Remaining sample was frozen and stored at –80°C. For this research study, these samples were thawed, aliquoted (40 ml), and flash frozen in liquid nitrogen, and stored at –80°C locally. Samples were anonymized and randomized so that researchers were blinded to each participant’s underlying condition during sample preparation and spectral preprocessing.

### Sample preparation for Raman spectroscopy

CSF samples were thawed in room temperature (RT) water and vortexed well. CSF salts were exchanged for double-distilled H_2_O (ddH_2_O) using a 10 kDa MWCO filter (Vivaspin) as described below. Filters were first washed thoroughly by two 4-minute spins in 500µl ddH_2_O at 12,000 x g in a tabletop microcentrifuge at 20°C. Filtrate, collected in the bottom compartment, was discarded. 40µl CSF was then added to the top part of the filter and diluted to 500µl with ddH_2_O and centrifuged for 10 min at 12,000 x g, 20°C. This process of dilution and filtration was repeated three times. The desalted CSF was then transferred to a fresh tube and vortexed well. 1µl of the desalted CSF was deposited onto an aluminium coated (1000Å) microscope slide (SUBSTRATA, ANGSTROM Engineering®) and allowed to dry in a small, vented container at RT for 30 min.

### Raman spectroscopy

A Renishaw inViaTM Qontor microscope system was used for Raman spectroscopy. Briefly, the Raman system was calibrated to the 520 cm^−1^ reference peak of the internal silicon substrate prior to each experiment. The charge-coupled device (CCD) detector and spectrometer slit areas were aligned using the auto align function and the laser spot was manually aligned to the centre of the crosshairs using the camera. The desalted dried CSF droplets were located and brought into focus using a Leica DM 2500-M bright field microscope and an automated 100 nm-encoded XYZ stage. Spectral data were collected, and parameters were determined using Renishaw WIRE5.5 software. The samples were excited using a 785nm laser at 50% power focused through a Leica 50x long working distance objective (numerical aperture = 0.5). Laser power at the sample was approximately 6.25 mW. Each Raman spectrum was collected for 30s, consisting of two 15s measurement acquisitions.

Thirty spectra were collected from the dried droplets. Only the bottom half of each droplet was measured, as seen through the camera. All raw spectra consisted of 1015 variables and detected using a Peltier cooled CCD (1024 pixels × 256 pixels) after dispersion through a 1800L/mm diffraction grating with a range of 659cm^−1^ –1761cm^−1^ with a resolution of 1.09cm^−1^. As multiple acquisitions were acquired per spectrum, cosmic rays were removed manually using the WIRE5.5 zap function after each spectral measurement. Background spectra from the aluminium surface were measured in 3 roughly evenly spaced locations around the measured region of the dried droplet using equivalent Z distances as for each sample measurement. All spectra were background-subtracted using an average of the three spatially independent spectra of the aluminium surface for each CSF droplet.

### Spectral preprocessing

Preprocessing and multivariate analysis were performed using the IRootLab plugin (0.15.07.09-v) for MATLAB® R2023a ^14^. High-frequency noise was removed using the Haar-wavelet denoising function with 6 decomposition levels. A fifth-order polynomial was used to remove fluorescence background. The ends of each spectrum were then anchored to the axis using the rubberband-like function. Spectral intensity normalization was applied using vector normalisation and spectra were standardized for outlier removal by principal component analysis (PCA). Identified spectral outliers were removed from the dataset for each patient. PCA was also applied at the level of patient resulting in the removal of 2 patient’s spectra from downstream analysis due to chemical contamination of sample (likely from blood during lumbar puncture).

### Spectral biomarker feature extraction

The Mann-Whitney U-Test per Raman shift of the spectrum was used to identify the variables that were most different between Non-AD and AD groups using all 3139 spectra in the training dataset. 10 spectral regions were identified, and the area of each region (A) was calculated to reduce the data from each region into a single variable. These 10 variables were used for clustering and classification (see classification). Chemical assignment of each region was based on the literature values. Linear discriminant analysis (LDA) was used for clustering non-AD and AD samples using all ten spectral biomarker values as input variables. LDA was performed using the IRootLab plugin (0.15.07.09-v) for MATLAB® R2023a ^14^.

### Classification

Samples remaining after the removal of outliers (n=141) were randomly split into a training set (113 patients, 80%) and a testing set (28 patients, 20%) whilst retaining class balance. This resulted in 3139 independent Raman spectra in the training dataset and 766 independent Raman spectra in the testing dataset. Model hyperparameters are shown in Supplementary Table 2. For classification of the complete Raman fingerprint, two different types of classification models were utilized; a convolutional neural network (CNN) and a linear support-vector machine (SVM) via the MATLAB® Classification Learner application. For extracted features (see spectral biomarker feature extraction) a bagged decision tree model was utilized. Feature reduction using PCA (for the CNN and SVM only) and misclassification costs were optimized manually through iterations, followed by hyperparameter tuning using default optimizable settings for each model. Specifically, 5 separate models were generated for the training dataset using a Bayesian optimization procedure of 20 iterations of hyperparameter optimization with ‘expected improvement per second plus’ for each of the models. The models were trained and validated using the training data and 5 k-fold cross validation. The best optimized model, of the 5 models, was then exported to MATLAB® to retrain a further 10 times, each time shuffling cross-validation groups, to assess model variability by SEM (standard error of the mean). Each of the 10 retrained models were evaluated on the unseen patient spectra (testing set) for classification of the unseen data into 2 distinct groups (Non-AD and AD). Classified spectra were grouped per patient to provide a classification score (%) for each patient based on the number of spectra that correctly classified the patient into AD or non-AD groups. Averaged scores were used to construct the final receiver-operating characteristic (ROC) curves to evaluate the robustness of the classification model generated.

### Feature importance

Global LIME (local interpretable model-agnostic explanations) and partial dependence analysis were performed using the MATLAB® Classification Learner application explain function. Global LIME was performed using all 3139 spectra in the training dataset using a tree model and a kernel width of 0.5. Partial dependence maxima were extracted from the highest score from the partial dependence plot for each feature.

### Partial correlation analysis

Partial correlation analysis was performed using IBM SPSS Statistics (v30) software. Adjustments were made for age and sex to find associations between classifier output scores and diagnostic scale of the patient’s PLM status (Paris-North Lille Montpellier biomarker scoring system), as well as between extracted features (spectral biomarkers) and log-transformed CSF biomarker levels, specifically Aβ42, total tau, and p-tau levels. The cutoffs for positive results used were: Aβ42 <680 pg/m (ptau >56 pg/ml, total tau >355 pg/ml). Total tau biomarker results were not available for 7 patients, so these patients were removed from partial correlation analysis for total tau. Two-sided *P* < 0.05 was considered statistically significant. We used False Discovery Rate (FDR) analysis to control for multiple comparisons for the extracted features and CSF biomarkers, accounting for 30 comparisons in total. Specifically, the Benjamini-Hochberg method was used with an FDR <5% considered statistically significant. Scatter plots were used to illustrate the correlations.

## RESULTS

### Subhead 1: Generation of an AD-specific Raman signature

We are interested in developing a rapid and sensitive differential diagnostic method for neurodegenerative diseases using biofluid Raman spectroscopy. The aim of the present study is to assess the accuracy at which Raman spectroscopy can classify Alzheimer’s disease (AD) from other conditions using CSF samples from a mixed clinical cohort. To address this, we measured Raman spectra of 143 CSF samples that were collected by lumbar puncture acquired from a cross-section of patients with neurological symptoms and stored in the University of Southampton Tissue Bank. The study cohort consisted of 67 AD patients and 76 non-AD patients (Table 1) who were diagnosed clinically by neurologists.

We have previously shown that proteins at physiological concentrations can be measured directly by Raman spectroscopy when samples are dried in droplets ≤1 µl using a method known as droplet deposition Raman spectroscopy (DDRS, ^15,16^). To utilize this method, salts and sugars were removed from CSF samples by centrifugal filtration before air drying on an aluminum surface in an array for DDRS. 30 Raman spectra were collected from each dried CSF droplet using a near-infrared laser (785 nm). Samples from up to 6 patients were thawed, desalted, dried and measured per experiment in a blinded fashion. The experimental and analytical workflow for the present study is depicted in Figure 1.

**Fig. 1.**
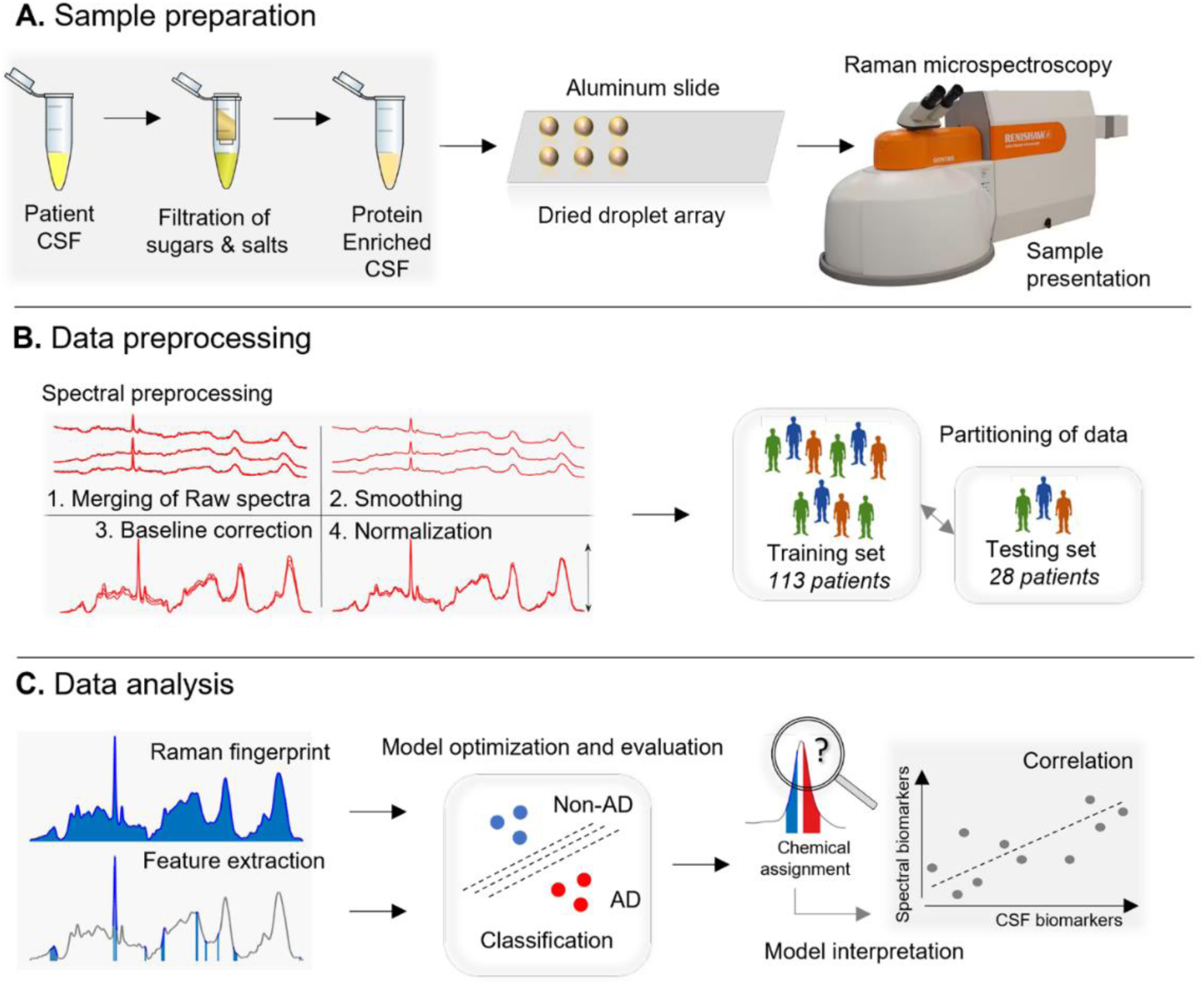
Schematic of workflow. **A**. Sample preparation for Raman spectroscopy. **B**. Preprocessing of Raman spectra for analysis. **C**. Analytical methods for classification of Raman spectra and identification of spectral biomarkers.

Raman spectra collected from CSF samples were preprocessed using established routines ^16–18^ and spectral outliers were removed using principal component analysis (PCA, see methods). The CSF Raman fingerprint is expectedly dominated by peaks that can be attributed to proteins (Figure 2A), including the characteristic phenylalanine ring-breathing mode at 1003 cm^−1^, the extended protein amide III region between 1200-1350 cm^−1^, and the protein amide I region between 1600-1700 cm^−1^. Interfering signals from salts and from sugars were removed by the filtration step and were subsequently undetectable by RS (Supplementary Figure 1). 2 CSF samples in the N=143 cohort were identified as outliers (Supplementary Figure 2) and these samples had notably darker colored solution, likely caused by blood contamination during lumbar puncture. Raman spectra from these 2 samples was subsequently excluded from the study analysis resulting in a final spectral dataset from 141 CSF samples.

**Fig. 2.**
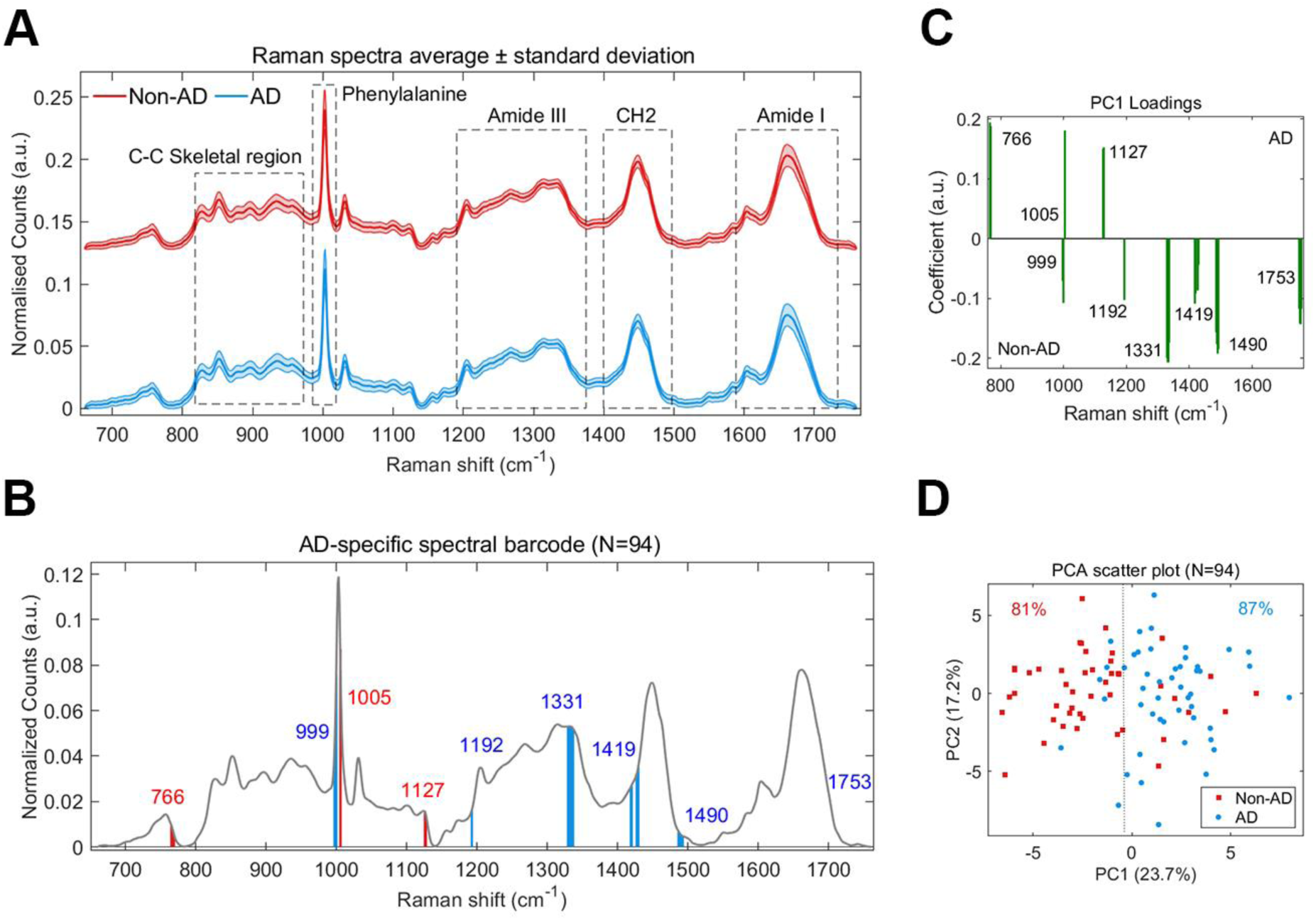
Generation of an AD-specific Raman signature. **A**. Average Raman spectra collected from CSF samples of 141 patients of whom 66 received a clinical diagnosis of Alzheimer’s disease (AD, blue trace) and 75 received a clinical diagnosis other than AD (non-AD, red trace). Key regions of the Raman fingerprint are annotated. **B**. Raman spectral barcode generated from identified spectral features using the Mann-Whitney U-test per Raman shift between high confidence non-AD (n=46) and AD spectra (n=48). The average reference spectrum is shown in grey and spectral biomarker regions are shown in red (increased area in AD mean spectrum relative to non-AD) and blue (decreased area in AD mean spectrum relative to non-AD). **C-D**. PCA analysis of spectral biomarkers for the N=94 sample dataset. The PCA loadings (**C**) and the scatter plot for the transformed data across the PC1 and PC2 axis (**D**) and are shown. A linear cutoff is depicted as a dotted line.

As the sample cohort was heterogeneous, we initially sought to generate a Raman signature that was specific to AD. To do this, we utilized the PLM scale ^19^ that is used to classify AD based on the presence of Aβ42, p-tau and total tau (A, T, N) biological markers. Positivity for all 3 biomarkers indicates very high, 2 biomarkers indicates high, 1 biomarker indicates low and 0 biomarkers indicates very low likelihood of AD. For the non-AD group we selected Raman spectra from samples with a PLM score of 0, and for the AD group we selected Raman spectra from CSF samples with a PLM score of 2-3 as well as amyloid positivity (A+). Amyloid positivity was selected because tauopathy samples that were in the CSF cohort may have positivity for T and N and therefore a PLM score of 2. This resulted in a total of 94 samples (non-AD = 46, AD = 48). To find Raman signals specific to AD, we utilized a simple feature selection method developed previously ^20^ to generate a ‘spectral barcode’ for AD (Figure 2B). We observed 9 regions of the Raman spectrum that were significantly different between the non-AD and AD spectra (*P* < 0.01). We applied PCA to these AD spectral barcodes and generated a loadings spectrum highlighting the weight/coefficient of region (Figure 2C) and a scatter plot depicting the clustering of transformed spectral barcodes across PC1 and PC2 axes (Figure 2D). Using a linear cutoff, 84% of the spectral barcodes were clustered with their correct class (non-AD or AD).

### Subhead 2: Interpretable AD classification using Raman spectral barcode

We next wanted to identify the regions of the Raman spectrum that were most important for AD classification in the complete heterogenous cohort (N=141), regardless of ATN biomarker status. We wanted to generate ‘spectral biomarkers’ to allow the assignment of chemical information and correlations with ATN biomarker concentrations. The Raman spectra were first randomly partitioned into a training set consisting of spectra from 80% of the CSF samples (113 samples, 3139 spectra), and a testing set consisting of spectra from 20% of the CSF samples (28 samples, 766 spectra). The training set was used for feature selection, as well as to train, validate, and optimize the ML model, whilst the testing set, unseen by the model during the optimization process, was used for model evaluation.

Using our spectral barcoding method, 10 spectral regions were extracted from the training data and are shown as bars on the Raman spectrum in Figure 3A. These regions are notably similar to those observed in the clean AD cohort in Figure 2, with some small shifts in frequency and area, as well as the addition of the 1370 cm^−1^ region. To create univariate values for each region/bar, area under the curve (A) was calculated for each spectral region, with red bars representing an increase in feature area in AD spectra relative to non-AD spectra, and blue bars representing a decrease in feature area in AD spectra relative to non-AD spectra. We named the features #1 - #10 and ranked them based on the statistical significance between non-AD and AD spectral areas, with feature 1 being most significant. Features 1 and 2, which are both related to the frequency of the Phenylalanine peak ∼1003 cm^−1^, were the most significant features between non-AD and AD groups and are shown in isolation for clarity in Figure 3B. We applied linear discriminant analysis (LDA) to these 10 spectral features (spectral biomarkers) to assess clustering of the 141 samples into AD and non-AD groups (Figure 3C). Using a linear cutoff, 21% overlap was observed between the two groups, with 77% of non-AD samples and 80% of AD samples correctly assigned.

**Fig. 3.**
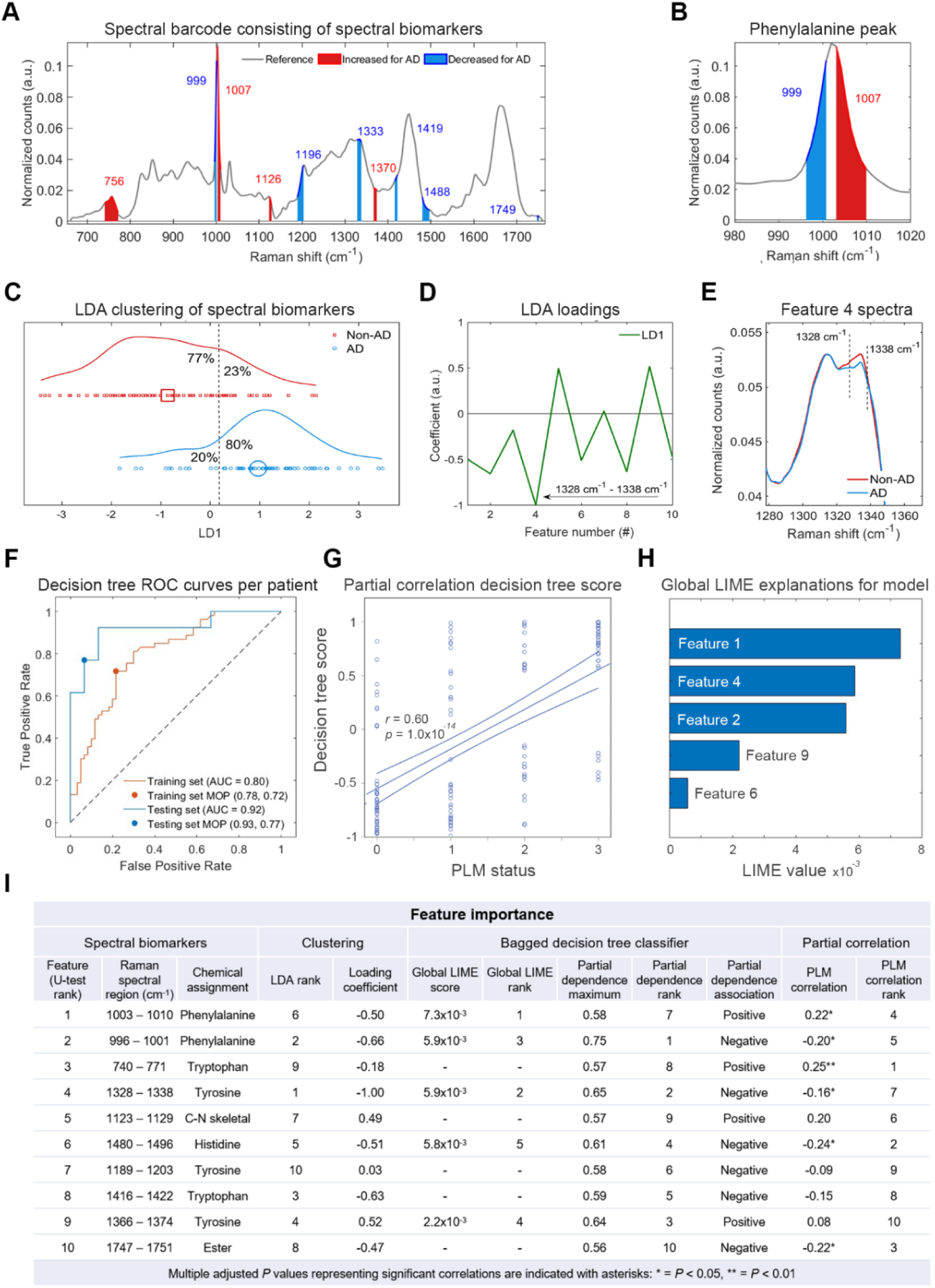
Interpretable AD classification using Raman spectral barcode. **A**. Raman spectral barcode generated from identified spectral features between non-AD and AD spectra using the training dataset. The average reference spectrum is shown in grey and spectral biomarker regions are shown in red (increased area in AD mean spectrum relative to non-AD) and blue (decreased area in AD mean spectrum relative to non-AD). **B**. Spectrum enhanced for clarity of the phenylalanine peak including feature 1 (red) and feature 2 (blue). **C-E**. Linear discriminant analysis (LDA) of spectral biomarkers for the complete N=141 sample dataset. The scatter plot for the transformed data across the LD1 axis (**C**) and the LDA loadings (**D**) are shown. Feature 4 is highlighted in the loadings (arrow) and the average Raman spectra for non-AD (red) and AD (blue) are shown (**E**). **F**. Receiver operating curve (ROC) analysis was performed for validation of the training data (orange curve) and evaluation of the testing data (blue curve). Raman spectra were separated into training and testing data at an 80:20 ratio at the level of patient. The bagged decision tree model was trained and optimized on the training data and evaluated on the testing data. Model operating points (MOP) representing sensitivity and specificity cutoffs for each model are depicted as a point on each curve. **G**. Partial correlation analysis of PLM biomarker status and Raman classification score for each patient CSF sample for the bagged decision tree model controlling for age and sex. 95% confidence intervals are depicted as minor lines on each side of the regression lines. **H**. Global LIME (local interpretable model-agnostic explanations) analysis to determine feature importance to model classification. **I**. Table summarizing each feature based on analyses. Adjusted P values are displayed after false detection rate (FDR) analysis for 10 correlations using the Benjamini-Hochberg method with FDR <5% considered statistically significant, *P < 0.05, **P < 0.01.

The LDA loadings in Figure 3D revealed that feature 4 A(1328 cm^−1^ −1338 cm^−1^) had the largest coefficient and most explained cluster assignment, with smaller contributions from all features except feature 7 A(1189 cm^−1^ −1203 cm^−1^). The spectral region for feature 4 is shown in Figure 3E and shows the difference between the average spectrum for non-AD samples (red) and AD samples (blue).

We next wanted to assess whether an interpretable classification model could be trained to classify AD in the testing group using only these 10 spectral biomarkers. To do this, we trained a bagged decision tree model for AD classification using all the spectra collected for each CSF sample in the training set after outlier removal to augment our data and improve model training ^12^. This was followed by the post-hoc binning of classification scores for each spectrum at the level of sample. ROC (receiver operating characteristics) curves and the area under the curves (AUC) were used to evaluate the efficacy of the models visually and quantitatively We observed acceptable accuracy for the optimized model for spectral classification (Supplementary Figure 4) for both the training data (AUC = 0.71) and the testing data (AUC = 0.79). Figure 3F shows the ROC curves constructed for the bagged decision tree when the spectra are binned at the level of sample for the training data (AUC = 0.80) and the testing data (AUC = 0.92), suggesting that optical biomarkers extracted from the training data are generalizable to the whole sample cohort. There was positive correlation between the decision tree classification score and PLM biomarker status for each patient (r = 0.60, p < 0.001) (Figure 3G and Table 2).

**Table 2.** Correlation for classification score per model corrected for age and sex.

|  |  |  |  |  |  |
| --- | --- | --- | --- | --- | --- |
| Model architecture | Model input variables | Correlations |  |  |  |
|  |  | PLM biomarker status | A biomarker concentration | T biomarker concentration | N biomarker concentration |
| CNN | PCs from Fingerprint | 0.64* | -0.47* | 0.55* | 0.41* |
| SVM | PCs from Fingerprint | 0.63* | -0.43* | 0.54* | 0.41* |
| Bagged decision tree | Extracted features | 0.60* | -0.42* | 0.40* | 0.33* |
| Footnotes: * Indicates corrected $P$ values representing significant correlations at $P < 0.001$, PLM (Paris-North Lille Montpellier biomarker scoring system), A (amyloid-β42), T (phosphorylated tau at site 181), N (total tau), CNN (convolutional neural network), SVM (support-vector machine) | | | | | |

We assessed the importance of spectral features of the decision tree model using multiple approaches. First, we applied global LIME (local interpretable model-agnostic explanations) analysis, which estimated that feature 1 A(1003 cm^−1^ - 1010 cm^−1^) assigned to phenylalanine ^21^, feature 4 A(1328 cm^−1^ - 1338 cm^−1^), assigned to tyrosine ^22^, and feature 2 A(996 cm^−1^ - 1001 cm^−1^) assigned to phenylalanine ^21^, contributed the most to AD classification based on an average local feature importance for all spectra (Figure 3H). This is in line with the initial statistical analysis used to generate the spectral barcode as features were numbered in order of statistical significance (features 1 - 10). We next calculated the partial dependence (PD) of each feature to show their individual effect on model prediction (Figure 3I). The top 3 features based on maximum PD score were feature 2 A(996 cm^−1^ - 1001 cm^−1^), feature 9 A(1366 cm^−1^ - 1374 cm^−1^), assigned to tyrosine ^22^, and feature 4 A(1328 cm^−1^ - 1338 cm^−1^). The partial dependency plot for each feature revealed a linear relationship between each spectral biomarker and prediction score (Supplementary Figure 4). For each method of analysis feature 2 A(996 cm^−1^ - 1001 cm^−1^), specifically arising from phenylalanine ring-breathing vibrations, was represented within the top 3 features for feature importance and is therefore likely to be important for prediction of AD. Feature 2 A(996 cm^−1^ - 1001 cm^−1^) also correlated with PLM status (r = −0.20, P = 0.03), although the strongest correlation was observed for feature 3 A(740 cm^−1^ −771 cm^−1^, r = 0.25, P = 0.009), which represents a skeletal bond vibration from tryptophan ^23^. Of the 10 features, 4 further features were correlated with PLM status, including feature 1 A(996 cm^−1^ - 1001 cm^−1^), feature 4 A(1328 cm^−1^ - 1338 cm^−1^), feature 6 A(1480 cm^−1^ - 1496 cm^−1^), assigned to histidine ^24^, and feature 10 A(1747 cm^−1^ - 1751 cm^−1^), assigned to esters ^25^.

Figure 4A shows the partial correlation matrix between the 10 extracted features (intensity area, A) and each of the 3 CSF biomarker concentrations (pg/ml) after adjusting for age and sex. Total tau biomarker results were not available for 7 patients, so these patients were removed from partial correlation analysis for total tau. After false discovery rate (FDR) correction using the Benjamini Hochberg adjustment, 4 significant associations were observed of the 30 tested. Feature 4 A(1328 cm^−1^ −1338 cm^−1^, tyrosine) negatively correlated with p-tau concentration (r = −0.299, P = 0.005), feature 3 A(740 cm^−1^ - 771 cm^−1^, tryptophan) positively correlated with p-tau concentration (r = 0.298, P = 0.005), and total tau concentration (r = 0.259, P = 0.023), and feature 6 A(1480 cm^−1^ - 1496 cm^−1^, histidine) positively correlated with Aβ42 concentration (r = 0.257, P = 0.020). Scatter plots depicting the correlations between CSF spectral biomarkers and CSF ATN markers are shown in Figure 4B-E respectively.

**Fig. 4.**
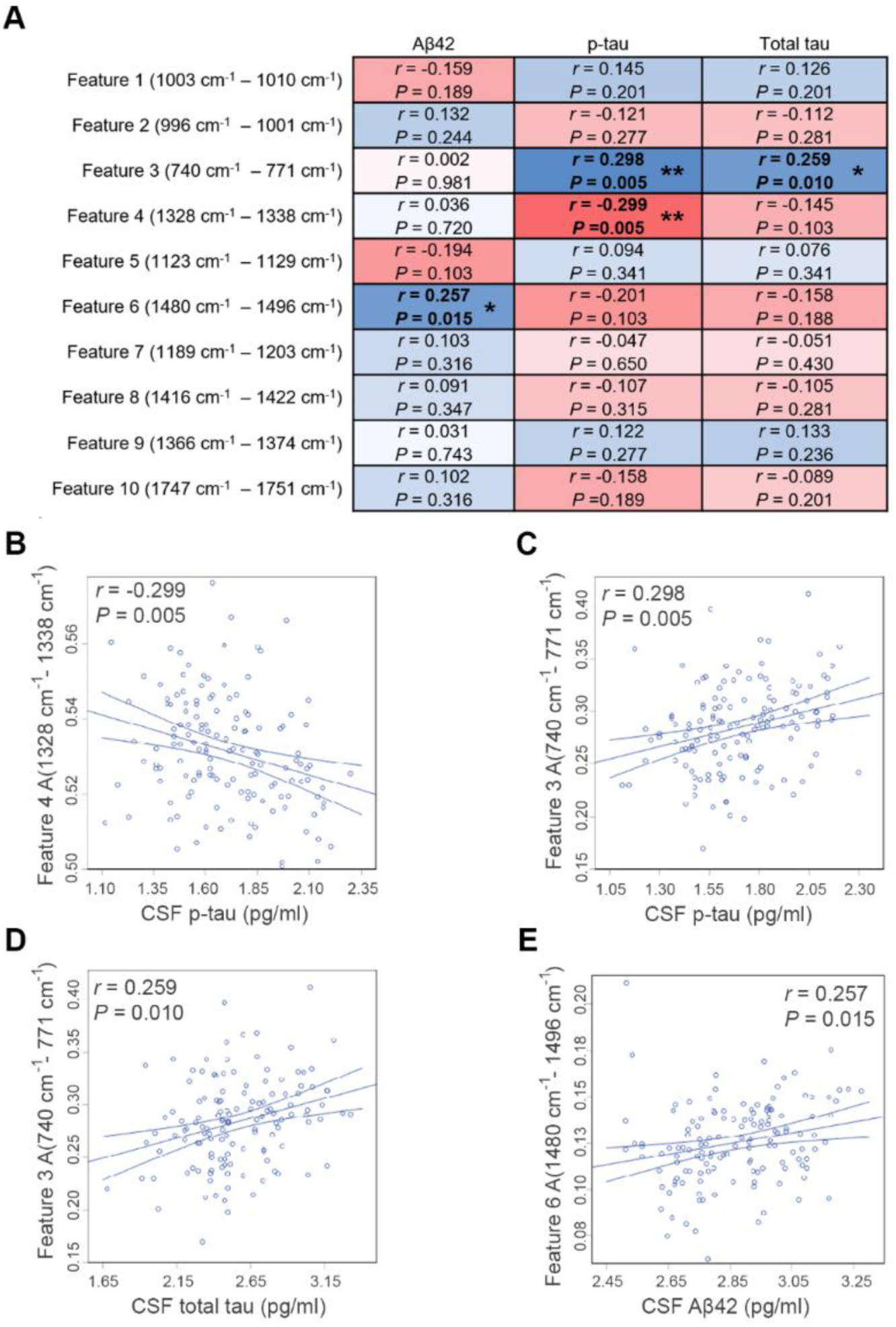
Correlation of spectral biomarker signals with ATN biomarker concentrations. **A**. Correlations between extracted Raman features (area within spectral range) and log transformed CSF antibody biomarker concentrations (Aβ42, p-tau, total tau). The partial correlation coefficients (r) were adjusted for age and sex. Adjusted P values are displayed after false detection rate (FDR) analysis for 30 correlations using the Benjamini-Hochberg method with FDR <5% considered statistically significant. **A.** Heatmap matrix for all 30 partial correlations displaying adjusted partial correlation coefficient (r) in blue for positive correlations and red for negative correlations, Adjusted P values are shown for each correlation: *P < 0.05, **P < 0.01. **B-E.** Scatter plots of extracted Raman features, and log transformed CSF antibody biomarker levels for statistically significant correlations.

### Subhead 3: AD classification using the whole Raman fingerprint

As the Raman fingerprint is a superposition of signals, that is, combined chemical information from many molecules, we next wanted to assess classification without limiting the spectral range. To reduce the feature space, we retained 95% of the variance extracted from the whole Raman fingerprint using PCA to train binary (AD vs non-AD) ML models with integrated feature selection algorithms to classify the spectra. We selected two different model architectures – support vector machine (SVM) and convolutional neural network (CNN). The SVM model is an all-purpose classifier that relies on data transformation using the kernel method before linear separation of classes, whilst the CNN can better learn unknown and complex features in datasets for classification but has a higher computational cost. Whilst neither method can be easily interpreted, they are each powerful in terms of ‘black box’ classification.

The ROC curves per spectrum for the optimized CNN and SVM models on the training set are shown in Supplementary Figure 5 (cross-validation: AUC for CNN = 0.79, for SVM = 0.75) and generalized well to the study cohort as demonstrated by the ROC curves for the testing set (evaluation: AUC for CNN = 0.79, for SVM = 0.73). The corresponding ROC curves for each sample in the training set is shown in Figure 5A (cross-validation: AUC for CNN = 0.96, for SVM = 0.94). Similar classification was observed for the samples in the testing set (evaluation: AUC for CNN = 0.92, for SVM = 0.93, Figure 5B). Each model demonstrated improved classification accuracy in comparison to the bagged decision tree model in Figure 3, but at the cost of model interpretability.

**Fig. 5.**
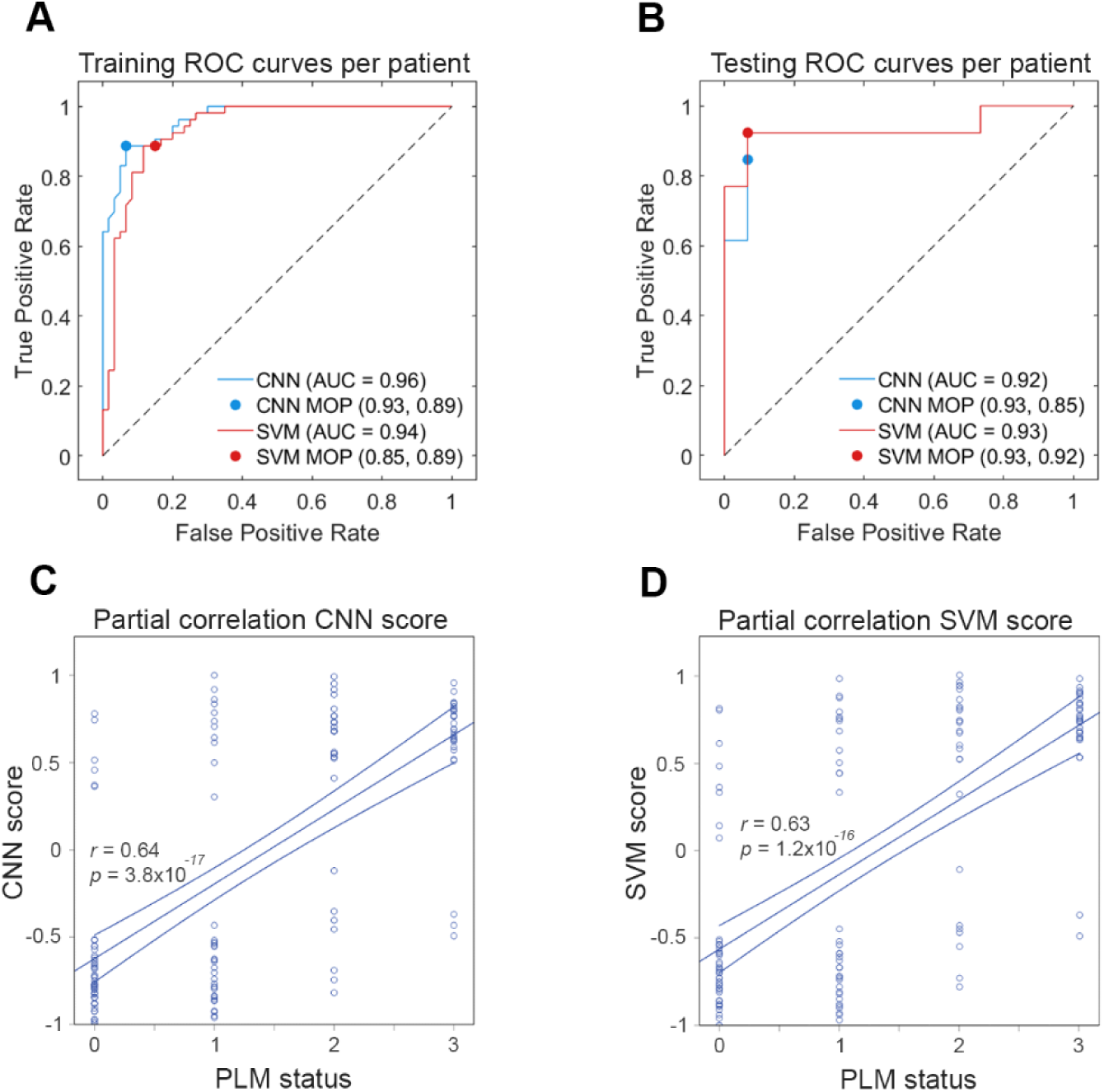
AD classification using the whole Raman fingerprint. **A-B**. Receiver operating curve (ROC) analysis was performed for validation of the training data (**A**) and evaluation of the testing data (**B**). Raman spectra were separated into training and testing data at an 80:20 ratio at the level of patient. CNN (blue traces) and SVM (red traces) models were trained and optimized on the training data and evaluated on the testing data. Model operating points (MOP) representing sensitivity and specificity cutoffs for each model are depicted as a point on each curve. D-E. Partial correlation analysis of PLM biomarker status and Raman classification score for each patient CSF sample for CNN (**C**) and SVM (**D**) models controlling for age and sex. 95% confidence intervals are depicted as minor lines on each side of the best fit line.

We wanted to understand how closely our CNN and SVM classification models corelated with Aβ42, p-tau and total tau (A, T, N) biological markers used for *ante-mortem* Alzheimer’s disease diagnosis. We controlled for age and sex in each case and observed a positive correlation between the CNN score and PLM biomarker status (r = 0.64, p < 0.001, Figure 5C), and between the SVM score and PLM biomarker status (r = 0.63, p < 0.001, Figure 5D), slightly higher than observed for the interpretable bagged decision tree model in Figure 3.

## DISCUSSION

In this study, we conducted Raman spectroscopy (RS) analysis of CSF samples from patients in a mixed clinical cohort. Our main aim was to assess the utility of RS for the classification of AD in a population with high clinical heterogeneity and thus reflecting the real-world. ML models were first trained to identify AD using Raman spectral fingerprints collected from 113 CSF samples and then evaluated on previously unseen spectra collected from 28 CSF samples. The optimized SVM model achieved 93% classification accuracy with 26 of 28 patients correctly classified as AD or non-AD during model evaluation. We also extracted features from the Raman fingerprint and used these as spectral biomarkers to develop an interpretable classification model for AD. These spectral biomarkers explained the majority, but not all, of the classification from the Raman fingerprint. Spectral biomarkers for AD primarily arose from protein-derived aromatic amino acid vibrations, the intensity of which directly correlated with biological CSF markers for AD, including Aβ42, phospho-tau 181 (p-tau), and total tau concentrations. Whilst these results support the utility of RS in aiding the clinical diagnosis of AD, larger cross-sectional studies are required to assess whether such high levels of discriminative accuracy are retained at a population level.

To date, two small proof-of-concept studies have used RS approaches to detect AD in human CSF. RS in combination with artificial neural networks and SVM discriminant analysis was used to identify AD with 84% accuracy in a cohort of 37 patients ^12^, whilst another study utilized a surface-enhanced variant of RS (SERS) in combination with a CNN to identify AD with 92% accuracy in a cohort of 19 patients ^11^. Despite small sample sizes, these studies provided initial evidence that AD signatures could be detected in CSF using RS. However, distinguishing AD from healthy controls is unlikely to be useful outside of population level screening, for which extracting CSF by lumbar puncture is not appropriate. CSF is often taken for ATN antibody biomarker analysis as part of the diagnostic pathway for AD ^3^ and RS has the potential to supplement such antibody markers and aid diagnostic decisions as demonstrated in this study. The advantages include; 1. RS is label-free and does not require antibodies or dyes, 2. RS requires very little sample with 40 µl CSF used for filtration and then 1 µl used for RS measurement, 3. RS measurement and classification can be automated and performed in seconds.

Larger proof-of-concept studies have demonstrated that RS can be used to detect AD from healthy controls in blood serum, N=56 ^26^, N=48 ^27^, and plasma, N=47 ^28, 29^. Although none of these studies met the following criteria: comparison to a gold standard of diagnosis, interpretable classification, a testing dataset for model evaluation, and n>20. Blood is a much more accessible biofluid for both diagnostics and screening and contains far higher protein levels (60-80mg/ml) than CSF (0.15-6mg/ml), also making it more amenable for RS analysis. It remains unknown whether RS can accurately classify AD using blood samples from a mixed clinical cohort as demonstrated for CSF in the present study. As well as limited clinical studies, a current barrier to the translation of RS is the size and cost of lasers and spectrometers. In the present study, we used a research-grade system, but system portability is possible ^30^, allowing large reductions in cost.

For the study cohort, the first limiting factor is sample size (n=143), which is relatively small considering the clinical heterogeneity observed within the population. Further to this, confirmation of AD diagnosis by post-mortem neuropathology was not possible as the work was conducted *ante-mortem*. Therefore, mixed pathologies cannot be ruled out and it is unknown how these may impact the Raman fingerprint. Importantly, rates of accuracy for clinical diagnosis of AD vary depending on disease stage and has been observed at 83% for patients with a clinical diagnosis of probable AD ^4^. In the present study, pathological ATN biomarkers and in many cases SPECT imaging were used to provide a robust clinical diagnosis for each patient to maximize diagnostic confidence. A subset of the sample cohort has also been classified in a previous research study (N=105) using AD-specific immunogenic inflammatory markers that correlated with total tau and p-tau levels ^31^, giving further confidence in clinical diagnosis. We also noted that the sex of the sample population was skewed towards male patients (65%). We addressed this by controlling for sex in all partial correlation analyses.

For the sample preparation, CSF samples were filtered using a microcentrifuge and spin filter to remove interference from sugars and salts. Whilst this simplified both measurement and analysis, such sample processing may hinder clinical translation, with cheaper and simpler solutions required such as one-step capillary separation ^32^. Whilst our correlation analyses suggest that variation in classification score has a molecular basis, it is possible that this can also be impacted by measurement heterogeneity. Much research effort is being directed towards the development of quality control or standards of practice for spectroscopic measurement pipelines ^33^.

For the analytical method, we required ‘black-box’ CNN and SVM classifiers for the most accurate classification of AD samples. These algorithms are prone to overfitting and their lack of interpretability reduces confidence in their application ^34^. To offset these problems, we further trained an interpretable classification model ^35^ based on features extracted from the Raman spectrum, i.e. spectral biomarkers. Whilst an increase in error rate was observed for classification, this approach enabled us to assign specific spectrochemical information to AD classification. Whilst these spectral biomarkers may not relate precisely to the features selected by the black-box classifiers, they provide an interpretable explanation of class identity. This enabled us to correlate spectral biomarkers to biological CSF markers resulting in 4 associations between 3 spectral biomarkers and all 3 CSF biomarkers (Aβ42, p-tau, and total tau). It is important to note that peaks in the Raman spectrum are not necessarily independent and can be correlated. They are a superposition or *fingerprint* of the vibrations arising from all molecules in a given mixture and are therefore akin to an ‘omic signature. We therefore propose that ‘spectromic’ signatures should be considered holistically during classification, and interpretable classification should be used to uncover associations with known biomarkers supporting mechanistic understanding hence enhancing confidence in classification.

Our study is the first to show the feasibility of applying the simple, holistic and label-free (unbiased) spectral biomarker approach using RS for dementia diagnosis on a real-world cohort. Importantly, samples from patients with other neurodegenerative diseases and long-term chronic conditions were clearly distinguished from patients clinically diagnosed with AD supporting the fact that RS provides a biochemical fingerprint, which will be disease-specific because the pathological processes are distinct in different diseases. We showed that Raman spectral biomarkers arising from protein-derived aromatic amino acids primarily explained spectral differences between the AD and non-AD CSF, likely due to proteomic changes specific to AD. Whilst the application of RS to dementia diagnosis remains a research field in relative infancy, our study demonstrates utility in a real-world population and suggests that there is potential for future clinical translation after larger and more diverse clinical studies. Finally, as this method is holistic and does not rely on a specific analyte, the full potential of Raman spectroscopy can be realized if spectral biomarkers are identified that are specific to other diseases that cause dementia including but not limited to vascular dementia (VaD), frontotemporal dementia (FTD) and dementia with Lewy bodies (DLB), to facilitate the stratification of clinically overlapping neurodegenerative diseases. Compared to current and emerging methods including blood-based biomarkers, the Raman spectroscopy approach neither requires sophisticated instrumentation or specialized labs nor reagents and can be implemented in a variety of clinical and healthcare settings without the need of skilled operators potentially offering unprecedented scalability and accessibility that can transform dementia diagnosis.

## List of Supplementary Materials

Fig. S1 Salts and sugars are removed from CSF after desalting

Fig. S2 Principal Component Analysis (PCA) for outlier analysis at the level of sample

Fig. S3 ROC curves per spectrum for CNN and SVM models

Fig. S4 ROC curves per spectral barcode for bagged decision tree model

Fig. S5 Partial dependence plots for bagged decision tree explaining AD

Table S1 Patient clinical diagnoses and CSF biomarker status

Table S2 Optimized model hyperparameters

## Data Availability

All data produced in the present study are available upon reasonable request to the authors

## Acknowledgments

The authors give thanks to Dr. Joe Chouhan and Professor Jessica Teeling for their support with sample preparation and the National Academy for Health Research (NIHR) Research Support Service for assistance with study design.

## Funding

Alzheimer’s Research UK (ARUK) Research Fellowship ARUK-RF2022B-010 (GD)

Engineering and Physical Sciences Research Council grant EP/T020997/1 (SM)

Engineering and Physical Sciences Research Council grant EP/V038036/1 (SM)

NIHR Southampton Biomedical Research Centre (BRC) support (CMK)

Institute for Life Sciences (IfLS) support (GD, AM, SM)

National Institute for Health Research (NIHR) Southampton Antimicrobial Resistance Laboratory support (SM)

## Author contributions

Conceptualization: GD, AM, CK, SM

Methodology: GD, SKM, LK, NH, AP, BG, AM, CK, SM

Investigation: GD Visualization: GD

Data curation: GD, SKM, AP

Funding acquisition: GD, AM, CK, SM Project administration: GD, SKM, AP Supervision: GD, SM, AM, CK, SM Writing – original draft: GD

Writing – review & editing: GD, SKM, LK, AM, CK, SM

## Data and materials availability

All processed data are available in the main text or the supplementary materials. Raw spectral data and code will be freely available on institutional repository upon manuscript publication.

## References and Notes

1. Lee JC, Kim SJ, Hong S, Kim Y. Diagnosis of Alzheimer’s disease utilizing amyloid and tau as fluid biomarkers. Exp Mol Med. May 9 2019;51doi:ARTN 53 10.1038/s12276-019-0250-2

2. Markesbery WR. Neuropathological criteria for the diagnosis of Alzheimer’s disease. Neurobiol Aging. Jul-Aug 1997;18(4):S13–S19. doi:Doi 10.1016/S0197-4580(97)00064-X

3. Jack Jr CR, Andrews JS, Beach TG, et al. Revised criteria for diagnosis and staging of Alzheimer’s disease: Alzheimer’s Association Workgroup. Alzheimers & Dementia. Aug 2024;20(8):5143–5169. doi:10.1002/alz.13859

4. Beach TG, Monsell SE, Phillips LE, Kukull W. Accuracy of the Clinical Diagnosis of Alzheimer Disease at National Institute on Aging Alzheimer Disease Centers, 2005-2010. J Neuropath Exp Neur. Apr 2012;71(4):266–273. doi:10.1097/NEN.0b013e31824b211b

5. van Dyck CH, Swanson CJ, Aisen P, et al. Lecanemab in Early Alzheimer’s Disease. New Engl J Med. Jan 5 2023;388(1):9–21. doi:10.1056/NEJMoa2212948

6. Barthelemy NR, Salvado G, Schindler SE, et al. Highly accurate blood test for Alzheimer’s disease is similar or superior to clinical cerebrospinal fluid tests. Nat Med. Apr 1 2024;30(3):1085–1095. doi:10.1038/s41591-024-02869-z

7. Hampel H, Nisticò R, Seyfried NT, et al. Omics sciences for systems biology in Alzheimer’s disease: State-of-the-art of the evidence. Ageing Res Rev. Aug 2021;69doi:ARTN 101346 10.1016/j.arr.2021.101346

8. Pontecorvo MJ, Devous MD, Navitsky M, et al. Relationships between flortaucipir PET tau binding and amyloid burden, clinical diagnosis, age and cognition. Brain. Mar 2017;140:748–763. doi:10.1093/brain/aww334

9. Devitt G, Howard K, Mudher A, Mahajan S. Raman Spectroscopy: An Emerging Tool in Neurodegenerative Disease Research and Diagnosis. Acs Chemical Neuroscience. Mar 2018;9(3):404–420. doi:10.1021/acschemneuro.7b00413

10. Kong K, Kendall C, Stone N, Notingher I. Raman spectroscopy for medical diagnostics - From in-vitro biofluid assays to in-vivo cancer detection. Adv Drug Deliver Rev. Jul 15 2015;89:121–134. doi:10.1016/j.addr.2015.03.009

11. Yu XK, Srivastava S, Huang S, Hayden EY, Teplow DB, Xie YH. The Feasibility of Early Alzheimer’s Disease Diagnosis Using a Neural Network Hybrid Platform. Biosensors-Basel. Sep 2022;12(9)doi:ARTN 753 10.3390/bios12090753

12. Ryzhikova E, Ralbovsky NM, Sikirzhytski V, et al. Raman spectroscopy and machine learning for biomedical applications: Alzheimer’s disease diagnosis based on the analysis of cerebrospinal fluid. Spectrochim Acta A Mol Biomol Spectrosc. Mar 5 2021;248:119188. doi:10.1016/j.saa.2020.119188

13. Keshavan A, Wellington H, Chen ZB, et al. Concordance of CSF measures of Alzheimer’s pathology with amyloid PET status in a preclinical cohort: A comparison of Lumipulse and established immunoassays. Alzh Dement-Dadm. 2020;12(1)doi:ARTN e12097 10.1002/dad2.12097

14. Trevisan J, Angelov PP, Scott AD, Carmichael PL, Martin FL. IRootLab: a free and open-source MATLAB toolbox for vibrational biospectroscopy data analysis. Bioinformatics. Apr 15 2013;29(8):1095–1097. doi:10.1093/bioinformatics/btt084

15. Kocisová E, Kuizová A, Procházka M. Analytical applications of droplet deposition Raman spectroscopy. Analyst. Jun 10 2024;149(12):3276–3287. doi:10.1039/d4an00336e

16. Devitt G, Rice W, Crisford A, Nandhakumar I, Mudher A, Mahajan S. Conformational Evolution of Molecular Signatures during Amyloidogenic Protein Aggregation. ACS Chemical Neuroscience. 2019/11/20 2019;10(11):4593–4611. doi:10.1021/acschemneuro.9b00451

17. Devitt G, Crisford A, Rice W, et al. Conformational fingerprinting of tau variants and strains by Raman spectroscopy. Article. Rsc Advances. Mar 2021;11(15):8899–8915. doi:10.1039/d1ra00870f

18. Devitt G, Johnson PB, Hanrahan N, et al. Mechanisms of SARS-CoV-2 Inactivation Using UVC Laser Radiation. Acs Photonics. Dec 26 2023;11(1):42–52. doi:10.1021/acsphotonics.3c00828

19. Lehmann S, Dumurgier J, Schraen S, et al. A diagnostic scale for Alzheimer’s disease based on cerebrospinal fluid biomarker profiles. Alzheimers Res Ther. 2014;6(3)doi:ARTN 38 10.1186/alzrt267

20. Devitt G, Hanrahan N, Ramirez M, Mudher A, Mahajan S. A Novel Spectral Barcoding and Classification Approach for Complex Biological Samples using Multi-excitation Raman Spectroscopy (MX-Raman). ChemRxiv. 2024;doi:doi:10.26434/chemrxiv-2024-x0lhh

21. Hernández B, Pflüger F, Kruglik SG, Ghomi M. Characteristic Raman lines of phenylalanine analyzed by a multiconformational approach. J Raman Spectrosc. Jun 2013;44(6):827–833. doi:10.1002/jrs.4290

22. Hernández B, Coïc YM, Pflüger F, Kruglik SG, Ghomi M. All characteristic Raman markers of tyrosine and tyrosinate originate from phenol ring fundamental vibrations. J Raman Spectrosc. Feb 2016;47(2):210–220. doi:10.1002/jrs.4776

23. Hirakawa AY, Nishimura Y, Matsumoto T, Nakanishi M, Tsuboi M. Characterization of a Few Raman Lines of Tryptophan. J Raman Spectrosc. 1978;7(5):282–287. doi:DOI 10.1002/jrs.1250070511

24. Takeuchi H. Raman structural markers of tryptophan and histidine side chains in proteins. Biopolymers. 2003;72(5):305–317. doi:10.1002/bip.10440

25. Rothschild KJ, Asher IM, Anastassakis E, Stanley HE. Raman spectroscopic evidence for two conformations of uncomplexed valinomycin in the solid state. Science. Oct 26 1973;182(4110):384–6. doi:10.1126/science.182.4110.384

26. Paraskevaidi M, Morais CLM, Halliwell DE, et al. Raman Spectroscopy to Diagnose Alzheimer’s Disease and Dementia with Lewy Bodies in Blood. ACS Chemical Neuroscience. 2018/11/21 2018;9(11):2786–2794. doi:10.1021/acschemneuro.8b00198

27. Ryzhikova E, Kazakov O, Halámková L, et al. Raman spectroscopy of blood serum for Alzheimer’s disease diagnostics: specificity relative to other types of dementia. J Biophotonics. Jul 2015;8(7):584–596. doi:10.1002/jbio.201400060

28. Carmona P, Molina M, Calero M, Bermejo-Pareja F, Martinez-Martin P, Toledanog A. Discrimination Analysis of Blood Plasma Associated with Alzheimer’s Disease Using Vibrational Spectroscopy. Journal of Alzheimers Disease. 2013 2013;34(4):911–920. doi:10.3233/jad-122041

29. Habartová L, Hrubesová K, Syslová K, et al. Blood-based molecular signature of Alzheimer’s disease via spectroscopy and metabolomics. Clin Biochem. Oct 2019;72:58–63. doi:10.1016/j.clinbiochem.2019.04.004

30. Hubbard TJE, Shore A, Stone N. Raman spectroscopy for rapid intra-operative margin analysis of surgically excised tumour specimens. Analyst. Nov 21 2019;144(22):6479–6496. doi:10.1039/c9an01163c

31. Michopoulou S, Prosser A, Kipps C, Dickson J, Guy M, Teeling J. Biomarkers of Inflammation Increase with Tau and Neurodegeneration but not with Amyloid-beta in a Heterogenous Clinical Cohort. J Alzheimers Dis. 2022;89(4):1303–1314. doi:10.3233/JAD-220523

32. Guo WJ, Hansson J, van der Wijngaart W. Synthetic Paper Separates Plasma from Whole Blood with Low Protein Loss. Anal Chem. May 5 2020;92(9):6194–6199. doi:10.1021/acs.analchem.0c01474

33. Guo SX, Popp J, Bocklitz T. Chemometric analysis in Raman spectroscopy from experimental design to machine learning-based modeling. Nat Protoc. Dec 2021;16(12):5426-+. doi:10.1038/s41596-021-00620-3

34. Wongvibulsin S, Wu KC, Zeger SL. Improving Clinical Translation of Machine Learning Approaches Through Clinician-Tailored Visual Displays of Black Box Algorithms: Development and Validation. Jmir Med Inf. Jun 2020;8(6)doi:ARTN e15791 10.2196/15791

35. Hassija V, Chamola V, Mahapatra A, et al. Interpreting Black-Box Models: A Review on Explainable Artificial Intelligence. Cogn Comput. Jan 2024;16(1):45–74. doi:10.1007/s12559-023-10179-8

