## Supporting information for "Classification of Alzheimer’s disease in a mixed clinical cohort using biofluid Raman spectroscopy"

**Supplementary figures**

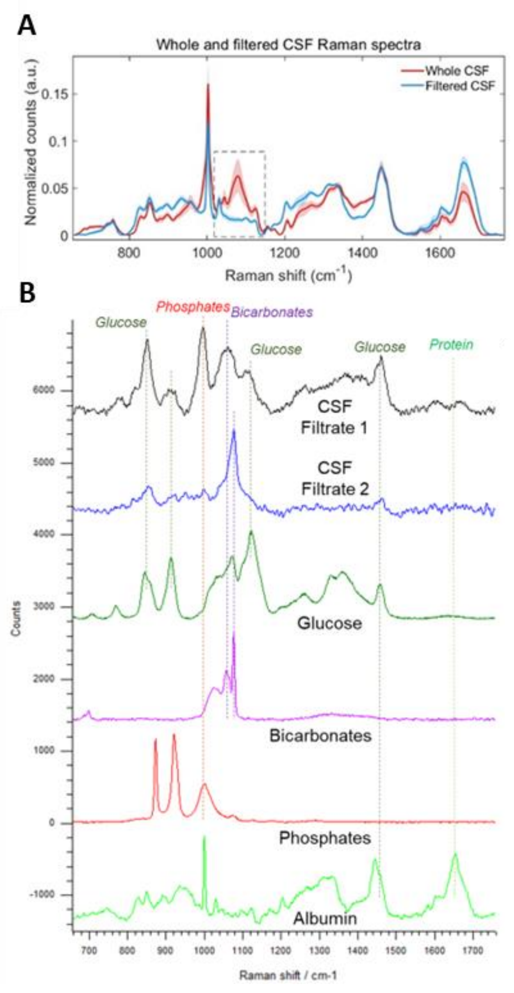


**Supplementary Figure 1. Salts and sugars are removed from CSF after desalting**
Raman spectroscopy analysis was used to determine that salts and sugars were not detectable after CSF filtration. Intra-sample measurement heterogeneity was reduced as a result. **A.** Average and standard deviation Raman spectra from whole CSF (red trace) and filtered CSF (blue trace) samples (n=98). **B.** Average Raman spectra from pure CSF components listed in order from top to bottom including CSF filtrate region 1 (black trace) and region 2 (blue trace), glucose (dark green trace), bicarbonate salts (pink trace), phosphate salts (red trace), and albumin (light green trace). Lines indicate aligning peaks between spectra from CSF filtrate and pure components.

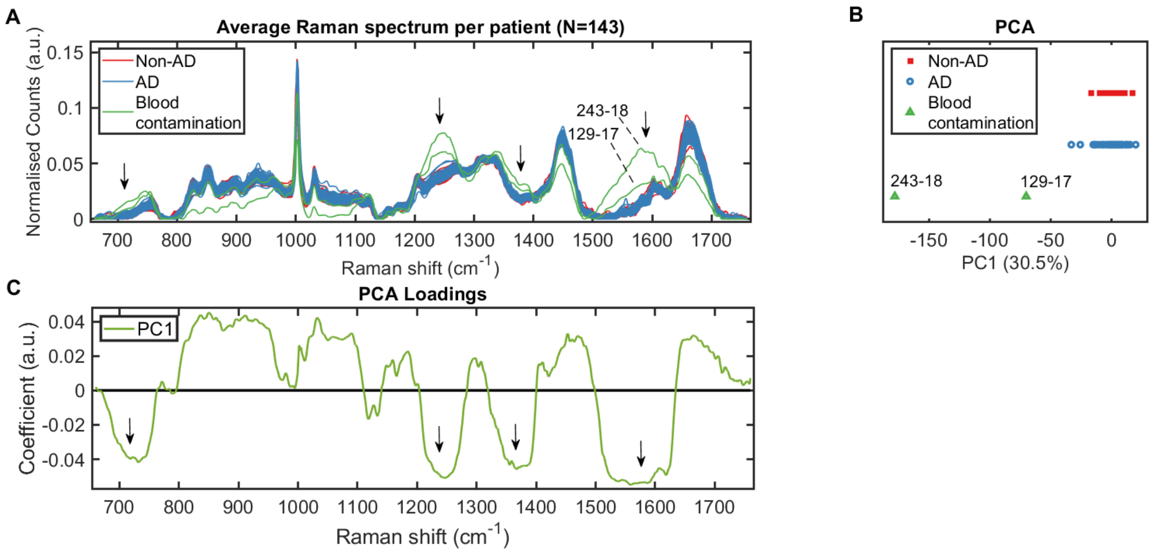


**Supplementary Figure 2. Principal Component Analysis (PCA) for outlier analysis at the level of sample**

Sample 243-18 (Non-AD) and sample 129-17 (AD) were removed during PCA outlier analysis. **A.** Raman fingerprint spectra represented as averages per sample for non-AD (red traces) and AD (blue traces) and outliers (green traces). Arrows indicate regions of highest difference for outlier spectra. **B.** The largest source of variance was identified by PCA (PC1). **C.** PCA loadings for PC1 showing spectral regions that explain variance across PC1 axis.

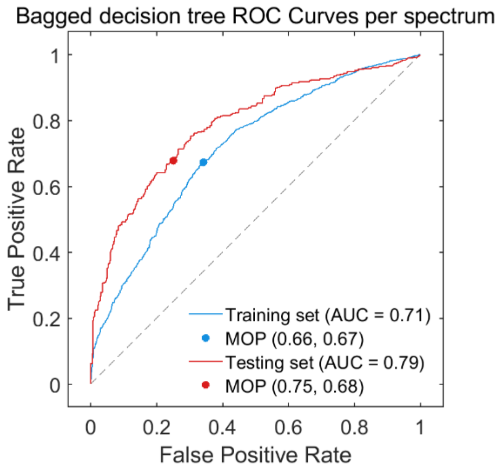


**Supplementary Figure 3 ROC curves per spectral barcode for bagged decision tree model**

Receiver operating curve (ROC) analysis was performed for validation of the training data (blue trace) and evaluation of the testing data (red trace). Raman spectra were separated into training and testing data at an 80:20 ratio at the level of patient. A bagged decision tree model was trained and optimized on the training data and evaluated on the testing data. Model operating points (MOP) representing sensitivity and specificity cutoffs for each model are depicted as a point on each curve.

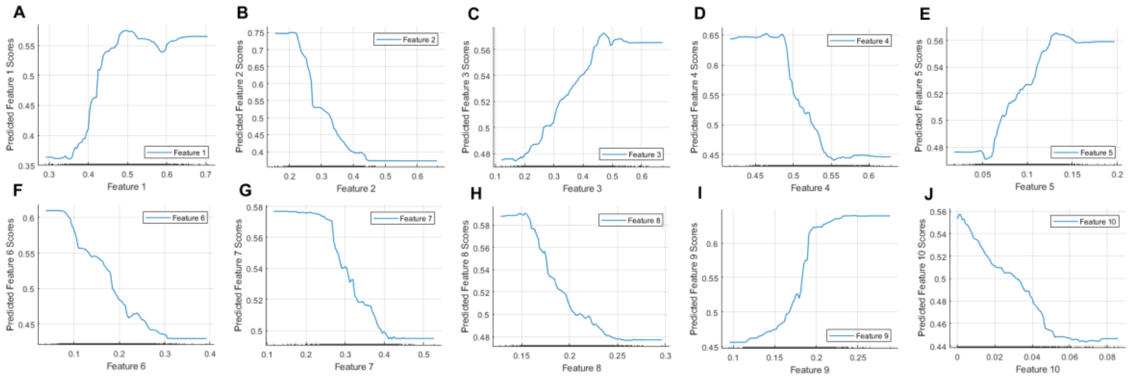


**Supplementary Figure 4. Partial dependence plots for bagged decision tree explaining AD**
Partial dependence plots for 10 extracted features/spectral biomarkers explaining AD for bagged tree classifier shown in Figure 3 of main manuscript; Feature 1 (**A**), feature 2 (**B**), feature 3 (**C**), feature 4 (**D**), feature 5 (**E**), feature 6 (**F**), feature 7 (**G**), feature 8 (**H**), feature 9 (**I**), feature 10 (**J**). All plots show a linear relationship between the given feature and classification score.

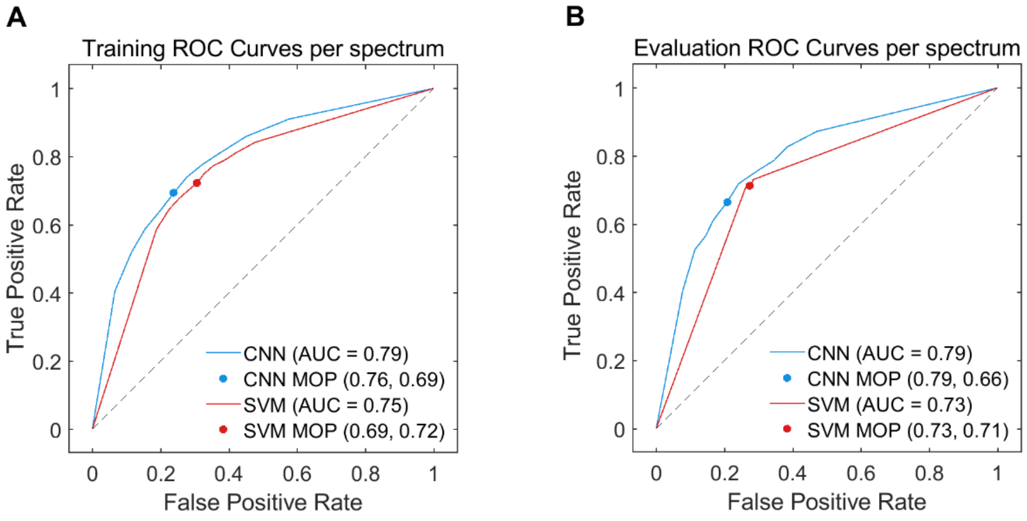


**Supplementary Figure 5 ROC curves per spectrum for CNN and SVM models**
Receiver operating curve (ROC) analysis was performed for validation of the training data (**A**) and evaluation of the testing data (**B**).  Raman spectra were separated into training and testing data at an 80:20 ratio at the level of patient. CNN (blue traces) and SVM (red traces) models were trained and optimized on the training data and evaluated on the testing data. Model operating points (MOP) representing sensitivity and specificity cutoffs for each model are depicted as a point on each curve.

| Supplementary Table 1. Patient clinical diagnoses and CSF biomarker status | | | | | |
|---|---|---|---|---|---|
| **Clinical diagnosis** | **Group** | **A** | **T** | **N** | **PLM** |
| Stable MCI | Non-AD | - | - | - | 0 |
| ?psychiatric | Non-AD | - | - | - | 0 |
| NPH | Non-AD | + | - | - | 1 |
| NPH | Non-AD | + | - | + | 2 |
| Subjective memory complaints (non-neurodegen) | Non-AD | - | - | - | 0 |
| Unlikely neurodegenerative | Non-AD | - | - | - | 0 |
| Functional neurological syndrome | Non-AD | - | + | - | 1 |
| ?NPH | Non-AD | - | - | - | 0 |
| Unlikely neurodegenerative | Non-AD | - | + | + | 2 |
| Likely vascular cognitive impairment | Non-AD | - | - | - | 0 |
| Possible NPH | Non-AD | - | - | - | 0 |
| PDD/DLB | Non-AD | + | - | - | 1 |
| Possible synucleinopathy | Non-AD | - | + | - | 1 |
| Likely Vascular cognitive impairment | Non-AD | - | - | - | 0 |
| Atypical FTD | Non-AD | + | - | - | 1 |
| Probable PSP | Non-AD | - | - | + | 1 |
| Vascular Cognitive Impairment | Non-AD | - | + | + | 2 |
| Complex partial seizures | Non-AD | - | - | - | 0 |
| DLB | Non-AD | - | - | - | 0 |
| Dementia plus seizures | Non-AD | + | - | - | 1 |
| ?NPH | Non-AD | - | - | - | 0 |
| NPH | Non-AD | + | - | - | 1 |
| Unclear (?lumbar compression / ?syncope) | Non-AD | - | - | + | 1 |
| NPH | Non-AD | + | - | - | 1 |
| CBD | Non-AD | + | - | - | 1 |
| Postural instability/dizziness | Non-AD | - | - | - | 0 |
| Migraine | Non-AD | - | - | - | 0 |
| ?Vascular disease /? Neurodegenerative / ?NPH | Non-AD | + | - | - | 1 |
| Subjective memory complaints (non-neurodegen) | Non-AD | - | - | + | 1 |
| Parkinsonism | Non-AD | - | - | + | 1 |
| FTD | Non-AD | - | - | + | 1 |
| Parkinsonism | Non-AD | - | - | - | 0 |
| idiopathic PD | Non-AD | - | - | - | 0 |
| NPH | Non-AD | + | - | - | 1 |
| Primary Progressive Aphasia (PPA-sv) | Non-AD | - | - | - | 0 |
| NPH | Non-AD | - | - | - | 0 |
| NPH | Non-AD | - | - | - | 0 |
| Depression | Non-AD | - | - | - | 0 |
| Psychiatric | Non-AD | - | + | + | 2 |
| NPH & Parkinson's disease | Non-AD | - | + | - | 1 |
| Likely vascular cognitive impairment | Non-AD | - | - | - | 0 |

| | | | | | |
|---|---|---|---|---|---|
| Multifactorial cognitive impairment, anxiety | Non-AD | - | - | - | 0 |
| PPA (FTD) | Non-AD | - | - | - | 0 |
| NPH | Non-AD | + | - | - | 1 |
| Possible bvFTD | Non-AD | - | - | - | 0 |
| Cognitive syndrome , anxiety depression | Non-AD | - | - | - | 0 |
| Ventriculomegaly, non reversible NPH | Non-AD | - | - | - | 0 |
| Cognitive Syndrome, non-neurodegenerative | Non-AD | - | - | - | 0 |
| PSP | Non-AD | + | - | - | 1 |
| Probable DLB, cerebrovascular | Non-AD | - | - | + | 1 |
| NPH, chronic headache, depression | Non-AD | - | - | - | 0 |
| Primary Progressive Aphasia (PPA-nf), progranulin mutation | Non-AD | - | - | - | 0 |
| NPH | Non-AD | - | - | - | 0 |
| Neurosarcoidosis, NPH | Non-AD | - | - | - | 0 |
| NPH | Non-AD | + | - | - | 1 |
| Seizures | Non-AD | - | - | - | 0 |
| PSP | Non-AD | - | - | - | 0 |
| DLB | Non-AD | + | - | - | 1 |
| NPH,Parkinsonism | Non-AD | + | - | - | 1 |
| Corticobasal syndrome | Non-AD | - | - | + | 1 |
| Secondary progressive multiple sclerosis (MS) | Non-AD | - | - | - | 0 |
| Non-AD cognitive decline | Non-AD | - | - | - | 0 |
| Non-AD cognitive decline | Non-AD | - | - | - | 0 |
| Primary Progressive Aphasia (PPA-sv) | Non-AD | - | - | - | 0 |
| NPH | Non-AD | - | - | - | 0 |
| Non-AD cognitive decline | Non-AD | - | - | - | 0 |
| Possible NPH | Non-AD | - | - | - | 0 |
| Frontemporal dementia | Non-AD | - | - | + | 1 |
| Cognitive decline – low mood | Non-AD | - | - | - | 0 |
| NPH | Non-AD | - | - | - | 0 |
| Frontotemporal dementia (behavioural variant) | Non-AD | - | - | + | 1 |
| DLB | Non-AD | - | - | - | 0 |
| DLB | Non-AD | - | - | - | 0 |
| NPH | Non-AD | + | - | - | 1 |
| Cortical dysplasia | Non-AD | - | + | - | 1 |
| AD | AD | + | - | + | 2 |
| AD / pituitary tumour | AD | + | - | - | 1 |
| Primary Progressive Aphasia | AD | - | - | - | 0 |
| Probable AD | AD | - | + | + | 2 |
| AD | AD | + | + | - | 2 |
| AD | AD | - | + | - | 1 |
| AD | AD | + | + | + | 3 |
| AD / Vascular | AD | - | - | - | 0 |

| | | | | | |
|---|---|---|---|---|---|
| AD | AD | + | + | + | 3 |
| Primary Progressive Aphasia ( PPA-nf) | AD | + | + | + | 3 |
| AD plus possible PD | AD | + | + | + | 3 |
| AD | AD | - | + | + | 2 |
| Posterior cortical atrophy | AD | - | - | + | 1 |
| AD | AD | + | + | + | 3 |
| AD | AD | + | + | + | 3 |
| AD | AD | + | + | + | 3 |
| AD | AD | + | + | + | 3 |
| AD | AD | + | + | + | 3 |
| AD | AD | + | + | + | 3 |
| AD | AD | + | + | + | 3 |
| AD | AD | + | + | + | 3 |
| AD | AD | + | + | + | 3 |
| Primary Progressive Aphasia (PPA) | AD | + | - | + | 2 |
| AD/LBD | AD | + | + | + | 3 |
| AD | AD | + | + | + | 3 |
| AD, epilepsy | AD | - | + | + | 2 |
| AD | AD | + | + | + | 3 |
| AD (logopenic) | AD | + | + | + | 3 |
| AD | AD | + | - | + | 2 |
| AD | AD | - | + | + | 2 |
| AD | AD | + | + | + | 3 |
| AD & NPH | AD | + | - | - | 1 |
| Possible AD | AD | - | - | - | 0 |
| Primary Progressive Aphasia (PPA-nf) | AD | - | - | - | 0 |
| Probable AD | AD | - | - | - | 0 |
| AD | AD | + | + | + | 3 |
| AD | AD | + | + | + | 3 |
| PPA (AD) | AD | + | + | + | 3 |
| AD | AD | - | + | + | 2 |
| Possible AD | AD | + | - | - | 1 |
| AD | AD | + | + | + | 3 |
| AD | AD | + | + | - | 2 |
| AD & NPH | AD | + | + | - | 2 |
| AD | AD | + | - | - | 1 |
| AD | AD | + | + | + | 3 |
| NPH & possible AD | AD | + | - | - | 1 |
| AD | AD | + | + | + | 3 |
| AD | AD | + | - | + | 2 |
| Possible AD | AD | + | - | - | 1 |
| AD | AD | + | + | + | 3 |
| AD | AD | - | + | + | 2 |

| | | | | | |
|---|---|---|---|---|---|
| Primary progressive aphasia (PPA-nf) | AD | + | - | + | 2 |
| AD | AD | + | + | - | 2 |
| Cognitive Impairment | AD | + | + | - | 2 |
| MCI | AD | + | + | - | 2 |
| AD | AD | + | + | - | 2 |
| AD | AD | + | + | - | 2 |
| AD | AD | + | + | - | 2 |
| Posterior cortical atrophy | AD | + | + | + | 3 |
| AD | AD | + | + | + | 3 |
| AD | AD | + | + | + | 3 |
| AD | AD | - | + | + | 2 |
| AD | AD | - | + | + | 2 |
| Posterior cortical atrophy (PCA) | AD | + | + | + | 3 |
| AD, Vascular Cognitive Impairment | AD | + | + | + | 3 |
| AD | AD | + | + | + | 3 |

Acronyms: A, (amyloid, Aβ42), T (tau, phosphorylated tau at residue 181), N (neurodegeneration, total tau), PLM (Paris-North Lille Montpellier scoring system), AD (Alzheimer's disease), MCI (mild cognitive impairment), NPH (Normal pressure hydrocephalus), Cl (cognitive impairment), PDD (Parkinson's disease dementia), DLB (dementia with Lewy bodies), FTD (frontotemporal dementia), PSP (progressive supranuclear palsy), CBD (corticobasal degeneration), MS (multiple sclerosis), bv (behavioural variant), NF-PPA (non-fluent primary progressive aphasia), svPPA (semantic variant primary progressive aphasia), LBD (Lewy body dementia), ? (low confidence in clinical diagnosis)

| Supplementary Table 2. Optimized model hyperparameters | |
|---|---|
| CNN model | |
| Number of fully connected layers | 2 |
| First layer size | 59 |
| Second layer size | 283 |
| Activation | ReLU |
| Standardize data | No |
| Regularization strength (Lambda) | $3.5605x10^{-9}$ |
| Misclassification costs | AD = 0.85, Non-AD = 1.00 |
| SVM model | |
| Kernel function | Linear |
| Kernal scale | Automatic |
| Multiclass coding | 1 vs. 1 |
| Standardize data | Yes |
| Box constraint level | 0.30809 |
| Misclassification costs | AD = 0.80, Non-AD = 1.00 |
| Decision tree model | |
| Ensemble method | Bag |
| Maximum number of splits | 88 |
| Number of learners | 324 |
| Number of predictors to sample | 8 |
| Standardize data | No |
| Misclassification costs | AD = 0.80, Non-AD = 1.00 |
